# Classifications of Logically Coherent Concurrent Diagnoses According to ICHD3: A Pilot Application of Automated Diagnosis Through Prime Representation

**DOI:** 10.1101/2022.08.21.22279042

**Authors:** Pengfei Zhang

**Author notes:** **Corresponding Author:** Pengfei Zhang, Department of Neurology, Robert Wood Johnson University Hospital - New Brunswick, New Jersey, United States.

## Abstract

**Introduction:** In clinical practice, headache presentations may fit more than one ICHD3 diagnoses. This project seeks to exhaustively list all these logically consistent “codiagnoses” according to ICHD3 criteria. We limit our project to cases where only two diagnoses are involved.

**Methods:** We included the ICHD3 criterias for “Migraine” (1.1, 1.2, 1.3), “Tension-type headache” (2.1, 2.2, 2.3, 2.4), “Trigeminal autonomic cephalalgias” (3.1, 3.2, 3.3, 3.4, 3.5), as well as all “Other primary headache disorders”. We excluded “Complications of migraine”(1.5) and “Episodic syndrome that may be associated with migraine” (1.6) since these diagnoses require codiagnoses of migraine as first assumption. We also excluded “probable” diagnosis criteria.

Each phenotype in the above criteria is assigned an unique prime number. We then encoded each ICHD3 criteria into integers, call “criteria representations”, through multiplication in a list format. “Codiagnoses representations” are generated by multiplying all possible pairings of criteria representations.

To eliminate logical inconsistent codiagnses, we manually encode a list of logically inconsistent phenotypes through multiplication: For example, headache lasting “seconds” would be logically inconsistent with “headache lasting hours”; the prime representation for both are multiplied together. We called this list the “inconsistency representations”.

All codiagnoses representation divisible by any inconsistency representations are filtered out, generating a list of codiagnoses represenation that are logically consistent. This list is then translated back into ICHD3 diagnoses.

**Results:** A total of 103 prime numbers were used to encode phenotypes from the included ICHD3 criteria diagnosis with 578 encodings generated. We generated 99 pairs of illogical phenotypes. Once illogical phenotypes were excluded, a total of 253,842 composite numbers representing unique dual-diagnosis clinical profiles were obtained. The number of profiles, although unique, yields duplicate dual diagnoses; once these duplicates are removed, we obtained 145 possible logical dual diagnoses.

Of the dual diagnoses, 2 contains with intersecting phenotypes due to subset relationships, 14 dual diagnoses with intersecting phenotype without subset relationships, 129 contains dual diagnoses as a result of non-intersecting phenotypes.

**Conclusion:** Prime number representations of primary headache disorders not only offer clinicians with an automated way of diagnosing headaches but also provides a powerful method of investigating co-diagnosis in headache classifications. Applications of this method to the investigations of dual diagnosis and headaches may offer insight into “loopholes” in the ICHD3 as well as potential explanation for sources of a number of controversies in headache disorders. Futures applications of the method includes extending the methodology to all of ICHD3.

## Introduction

In clinical practice, patient’s headache profiles may satisfy more than one ICHD3 criteria. These “co-diagnoses” can be a source of diagnostic challenges. Consider the cases where headache differential diagnosis lie between cluster headache and paroxysmal hemicrania when indomethacin trial cannot be attempted. In these cases the management maybe quite challenging.^1, 2^ A similar diagnostic dilemma exists between cluster headache and migraine with aura: Should we consider cluster headaches patients who exhibits aura symptoms a special subtype of cluster headache? Perhaps special subtype of migraine with aura? Or do these cases necessarily imply the co-existence of two separate headache disorders? (See de Coo et al and Silberstein et al versus et al. Peng et al)^3-7^ Finally, consider cases of identifiable date of onset of intractable headache without light or sound sensitivity: Should we consider these new daily persistent headaches or chronic tension type headaches? (Lobo et al vs. Robbins et al.) ^8,9^ The reader may have ready answer for each of the scenarios above; however, one has to agree that those opinions are not likely shared among our colleagues. It is not the goal of this paper is not to weigh in on each of these issues, but rather to identify the conditions of possibilities of their existence which arises as a result of our current classification system.

We can formulate these dilemma as the following: In the ICHD3 criteria, what are the headache disorders that can be logically diagnosed together? Or equivalently: Which headache presentations require more than one ICHD3 diagnoses to fully account for the clinical description? These sorts of questions can be properly called the “co-diagnosis” problems. (When co-diagnosis is restricted to two diagnoses, we use the term “dual diagnosis”.) The answer to the co-diagnosis problem is fundamental to headache classification as a scientific endeavor – it forces us to examine the limitation of our classification project and asks when the current paradigm is unable to assign unique diseases with unique diagnosis.

We propose that the solution to the co-diagnosis problem as linked to a novel technique of automated diagnosis through prime number representation: By encoding headaches phenotypes as prime numbers, and therefore diagnostic criteria as composite numbers, one can automate diagnosis of headache disorders through modular arithmetic. (This method was first presented at American Headache Society’s 2022 Denver Scientific Meeting.^10^) In this project we demonstrate that by encoding ICHD3 as numerical data it is possible to enumerate all possible dual diagnosis for primary headache disorders in ICHD3.

## Methods

There are two phases to this study. Since our technique is novel, the first phase will provide an intuitive description and rationale of our methods. Phase 2 then present an implementation for calculating ICHD3 dual diagnoses of primary headache disorders. Finally we analyses the result based on a paradigm of “intersecting phenotypes” outlined below.

### Phase 1: Automated Diagnosis Method’s Theoretical Construction

We will first demonstrate that any ICHD3 criteria can be readily translated into numerical data through prime number representations. Let us consider as an example the migraine criteria taken from ICHD3 below. (The demonstration here is an adaptation of a more formal mathematical treatment that is available as a preprint.^11^) We have excerpted the formal mathematical description as well as proof in the supplementary section to this paper. (*Not available in this preprint. Please refer to citation 11*.)

ICHD3 Criteria for Migraine without aura:

A. At least five attacks fulfilling criteria B–D
B. Headache attacks lasting 4–72 hours (when untreated or unsuccessfully treated)
C. Headache has at least two of the following four characteristics
  1. unilateral location
  2. pulsating quality
  3. moderate or severe pain intensity
  4. aggravation by or causing avoidance of routine physical activity (e.g. walking or climbing stairs)
D. During headache at least one of the following:
  1. nausea and/or vomiting
  2. photophobia and phonophobia
  3. Not better accounted for by another ICHD-3d diagnosis

Since all criteria are logical statements in disguise, one can translate the above into the following propositional logic statement: (We use the alphanumeric designation of the criteria as short-hand for each phenotype.)

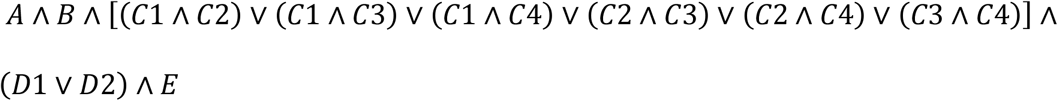

Now any logical statement can be translated into its disjunctive normal the form. (The disjunctive normative is simply a series of logic AND statements which are connected by OR.)^23^ Therefore one can translate the above into the following equivalent logical statement:

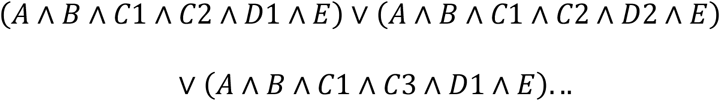

Given criteria in disjunctive normal form, we propose the following algorithm:

Step 1: Each phenotype in the ICHD3 is assigned a unique prime number. (Table 1) Negations crucial to diagnostic criteria are also assigned a prime number.

**Table 1.**
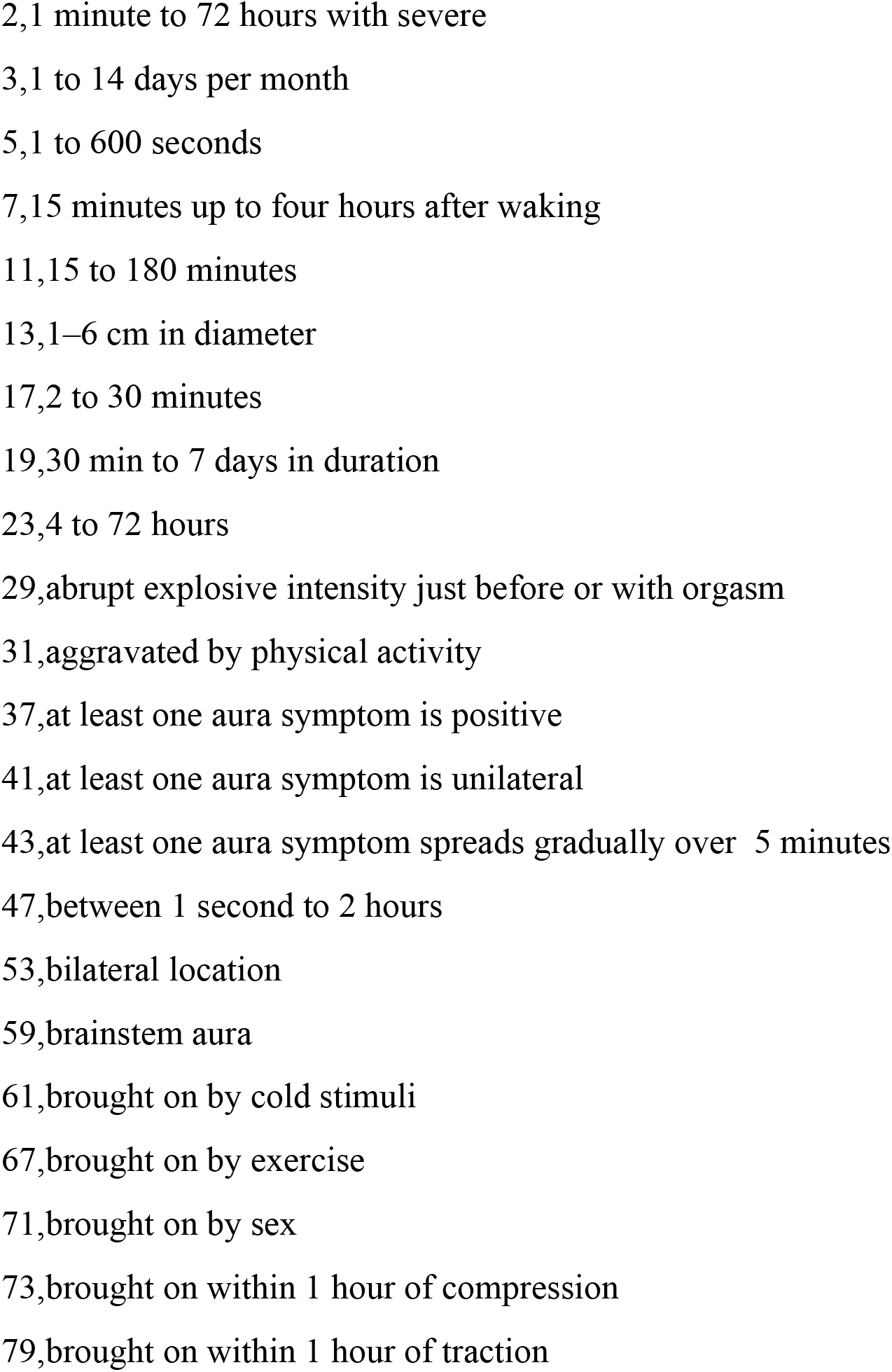
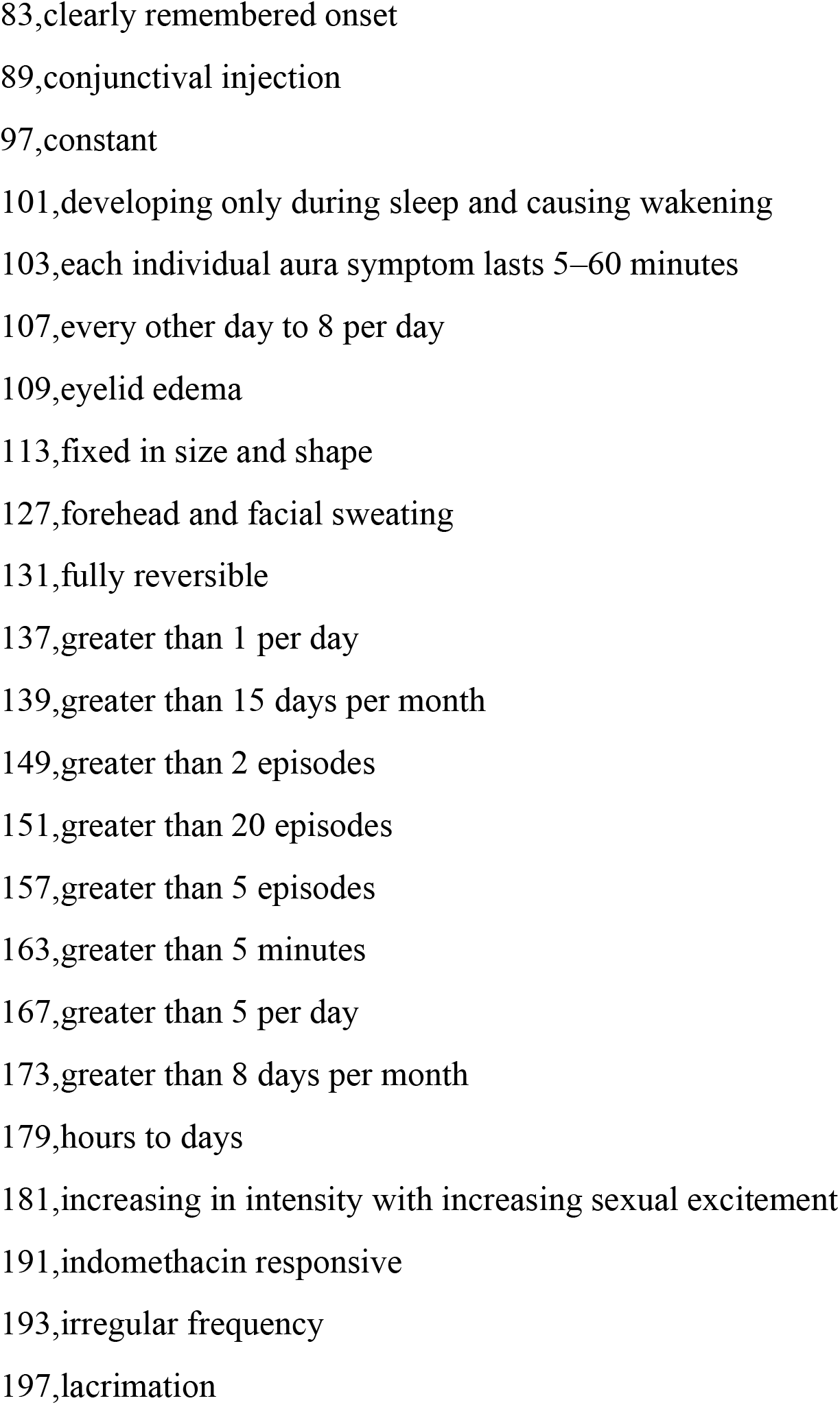
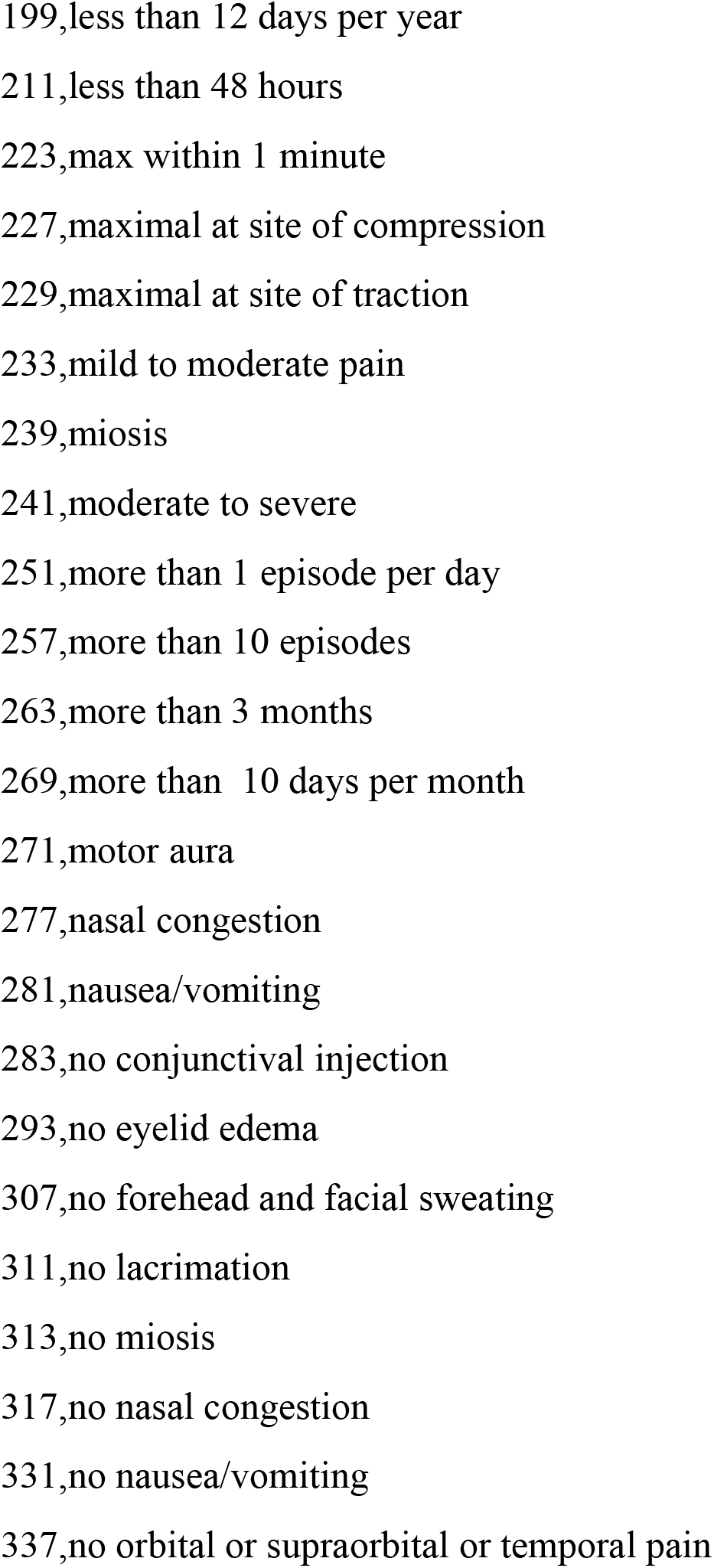
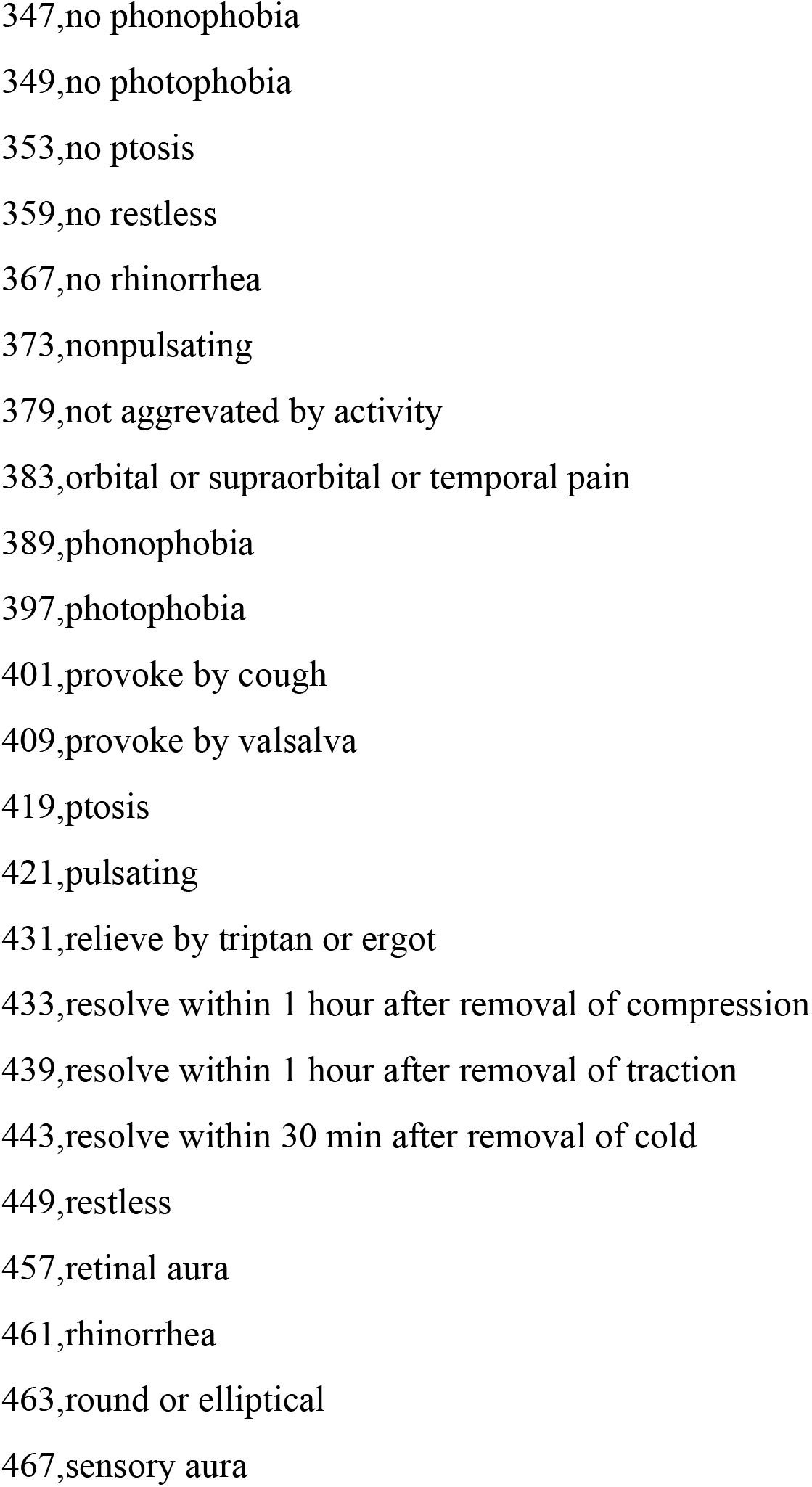
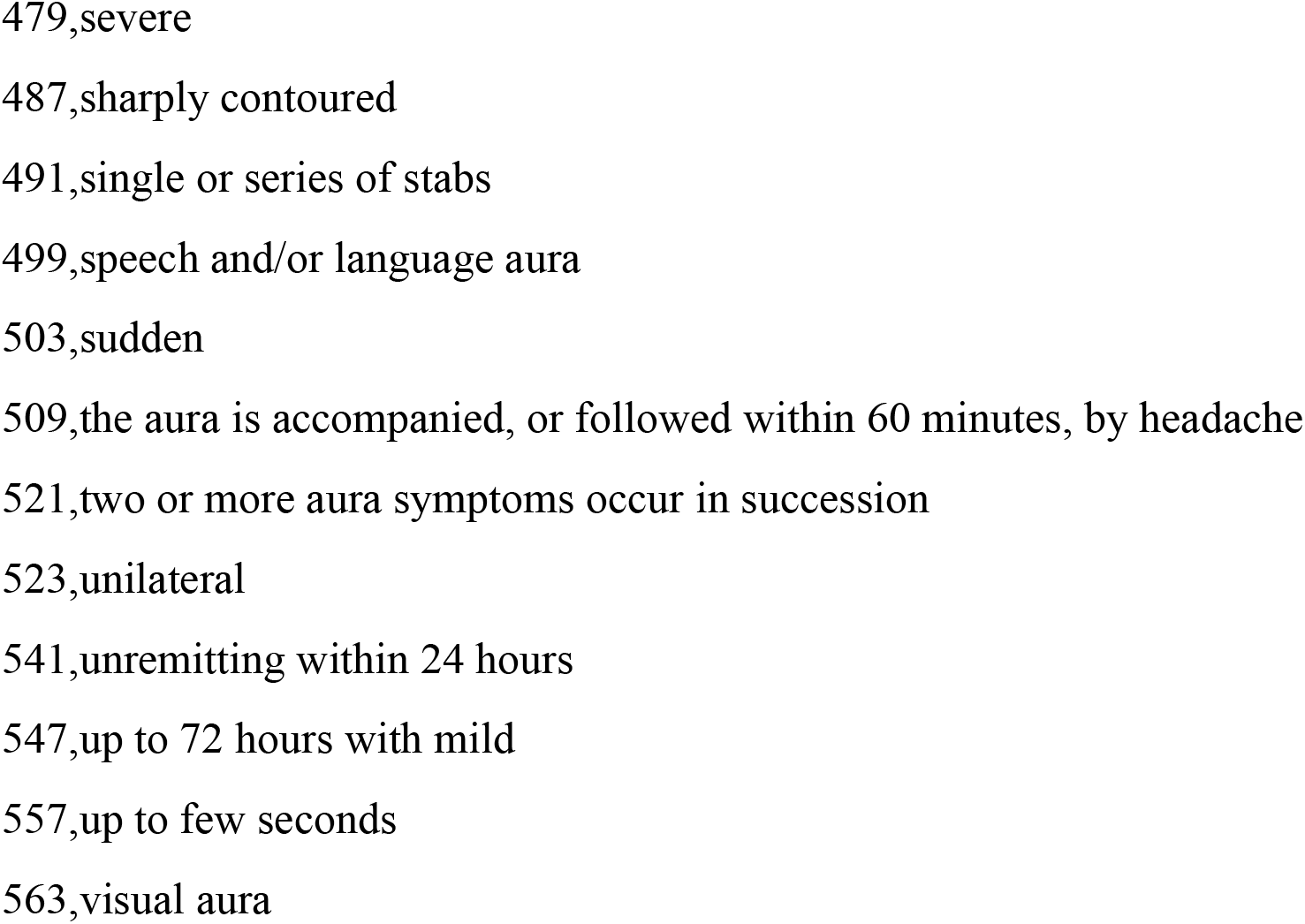
Sample Prime Number Assignments for Headache Phenotypes:

Step 2: If AND is used between two phenotypes, then the corresponding prime number for those two phenotypes are multiplied together. These are called encodings.

Step 3: All encodings that are separated by OR in the disjunctive normative form are then combined in a list.

Step 4: We do not encode the ubiquitous criteria “Not better accounted for by another ICHD-3d diagnosis”, given that it is recursively referring to the totality of ICHD3 encodings, creating a logical impasse.

For example, in migraine without aura the first conjunction in the distinctive normative form, when excluding criteria E, is *A* ∧ *B* ∧ *C*1 ∧ *C*2 ∧ *D*1. This is therefore encoding as:

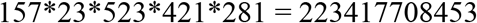

Repeating this procedure for totality of migraine without aura yields:

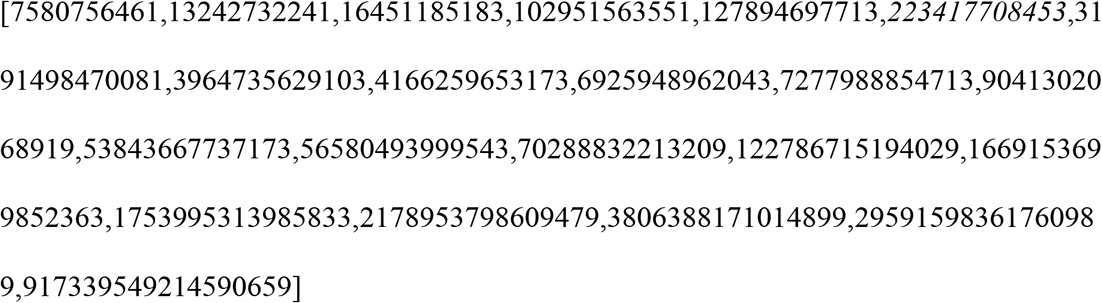

Now a patient’s headache profile, thought of as a collection of headache phenotypes, can be expressed using only logic conjunction. Therefore, a patient profile can be expressed as one composite integer. For example, a patient who has the phenotype of five headaches, each lasting 4 to 72 hours, unilateral, pulsating, with nausea, and photophobia can be represented as 157*23*523*421*281*397 = 88696830255841.

We now observe that a patient profile, represented as a composite number, must divide without remainder at least one number in its corresponding diagnosis’ encoding, assuming that a diagnosis exists. This observation forms the basis for automated diagnosis. For example, a patient profile having migraine without aura (such as 88696830255841 above) must have an encoding which divide by at least one number in the migraine without aura encodings (in this case, 223417708453) without remainder.

How this method works can be understood intuitively: Consider a patient profile satisfying migraine diagnosis. Then this patient’s profile, when translated into propositional logic, must satisfy one of the many conjunctions in migraine criteria’s disjunctive normative forms. (Since that is the implication of OR operator.) Encoding both the patients profile and the conjunction that it satisfies using our algorithm implies that both must share at minimum a collection of prime numbers. Since both sets of prime numbers are “bundled” together by multiplication in our algorithm, one of the two encodings must divide the other without remainder. The bigger of the two encoding must be the patient profile, since it has more “variable” to encode than those in the criteria. Therefore the patient encoding must be divisible by the encoding of one of the conjunction in the disjunctive normal form. We use prime number as building block for encoding since they have the property of not being able to divide by each other.

A caveat: although theoretically possible, in practice logical conflicts cannot exist within a patient’s clinical profile unless there exists more than one headache diagnosis – for example, it makes no sense to be both photophobic and not photophobic at the same time in either the criteria or user encoding.

Armed with the prime number automated diagnosis above, we can now tackle the uniqueness problem as follows: To obtain all possible combinations of dual diagnoses, we multiply all encodings of each ICHD3 diagnosis by those from another. This list, let’s call it M, represents patient profiles which satisfies two ICHD3 diagnoses concurrently. Now not every pairing of two ICHD3 encodings is possible as some of these pairing would contain phenotypes that are contradictory. This contradiction is manifested by the co-occurrence of two logically contradictory phenotypes in the same patient profile - if A and B are logically contradictory phenotypes, then its encoding is simply the prime representation of A multiplied by the prime representation of B. Therefore to obtain a non contradictory listing of duo diagnosis, we simply eliminate from the list M those that are divisible by A*B.

### Phase 2: Implementation of Automated Diagnosis for the Purpose of Extracting Dual Diagnoses

Due to the limitations in computational power, we only included ICHD3 primary diagnosis up to two layers deep in terms of ICHD3 classification. That is, ICHD3 criterias for “Migraine” (1.1, 1.2, 1.3), “Tension-type headache” (2.1, 2.2, 2.3, 2.4), “Trigeminal autonomic cephalalgias” (3.1, 3.2, 3.3, 3.4, 3.5), as well as all “Other primary headache disorders” are included in this study. We excluded “Complications of migraine”(1.5) and “Episodic syndrome that may be associated with migraine” (1.6) since these diagnoses require diagnoses of migraine as first assumption. We also excluded “probable” diagnosis criteria.

We then encoded all included criteria above into their numerical encodings. (Table 2) All possible take-two combinations encodings in Table 2 are multiplied together. We call this list M’ which represents the list M that was described in the last section.

**Table 2.**
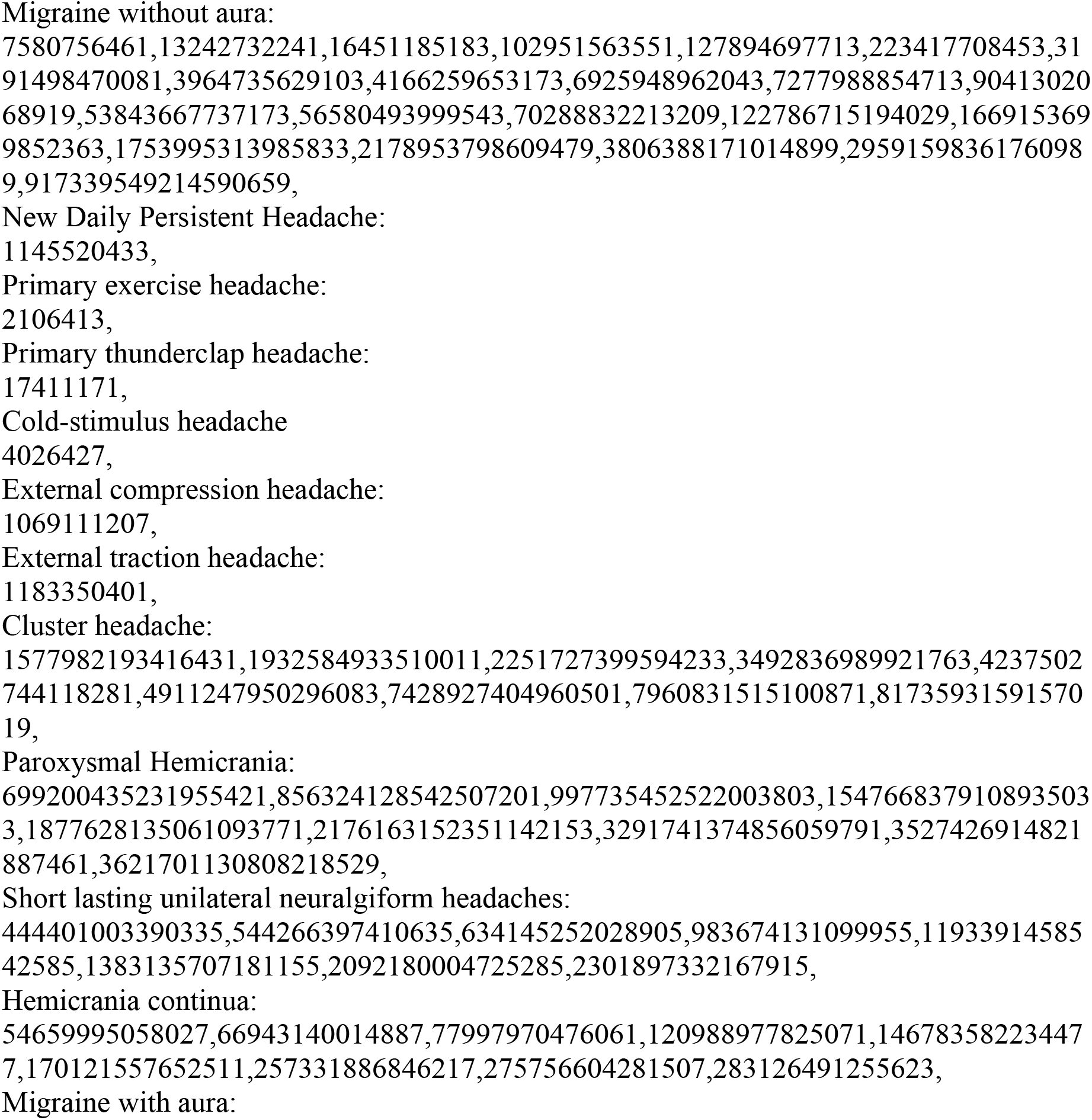
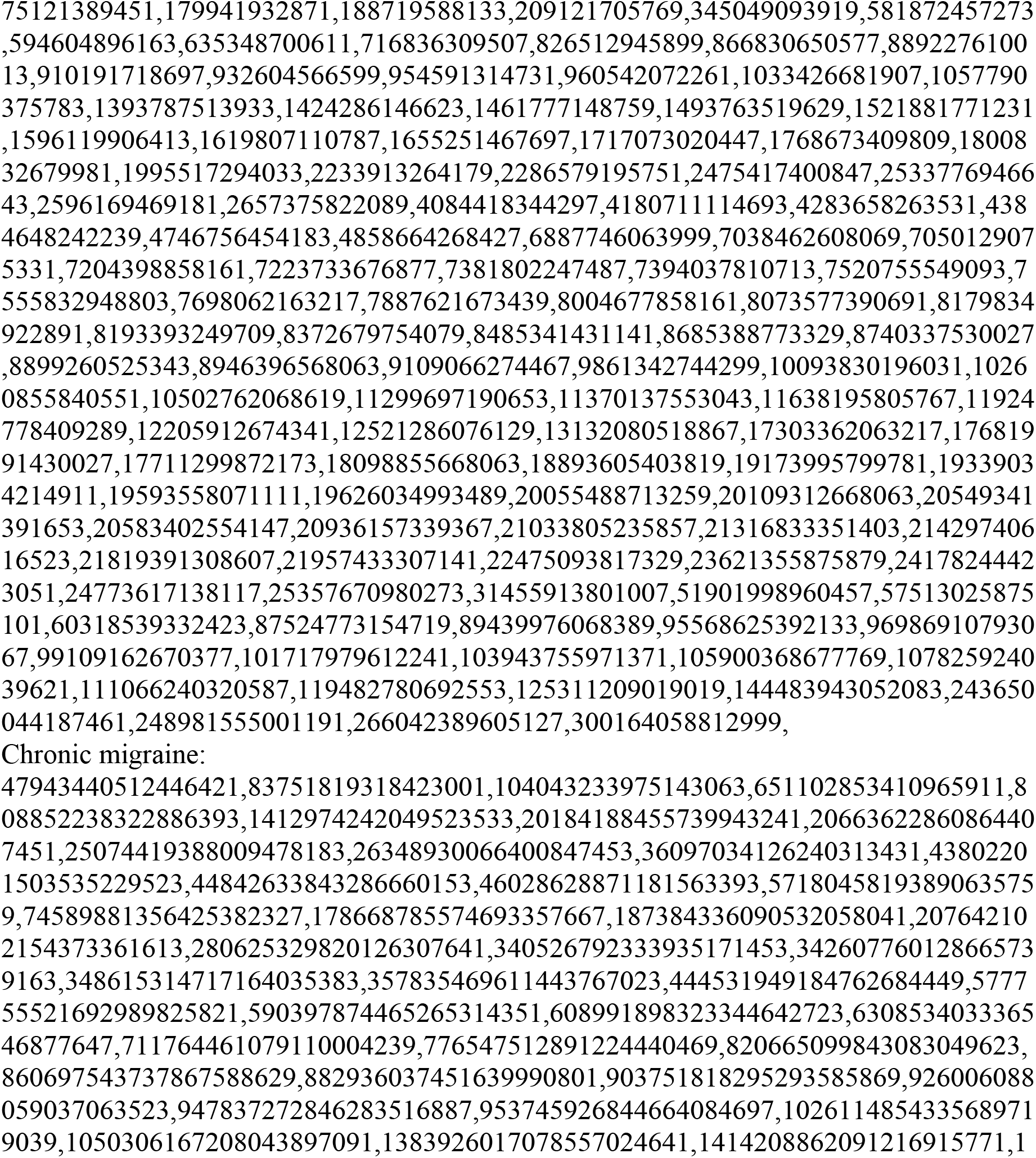
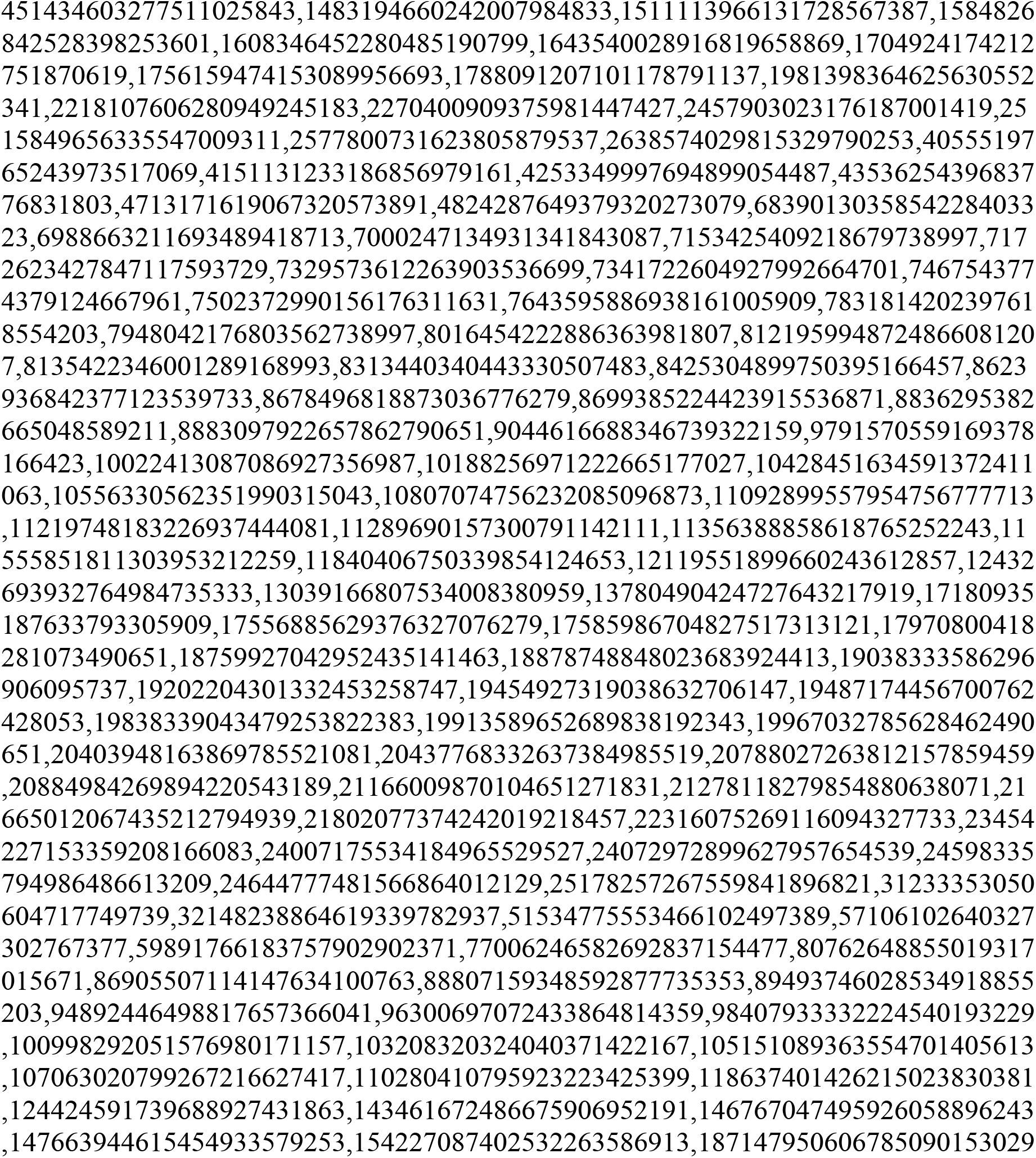
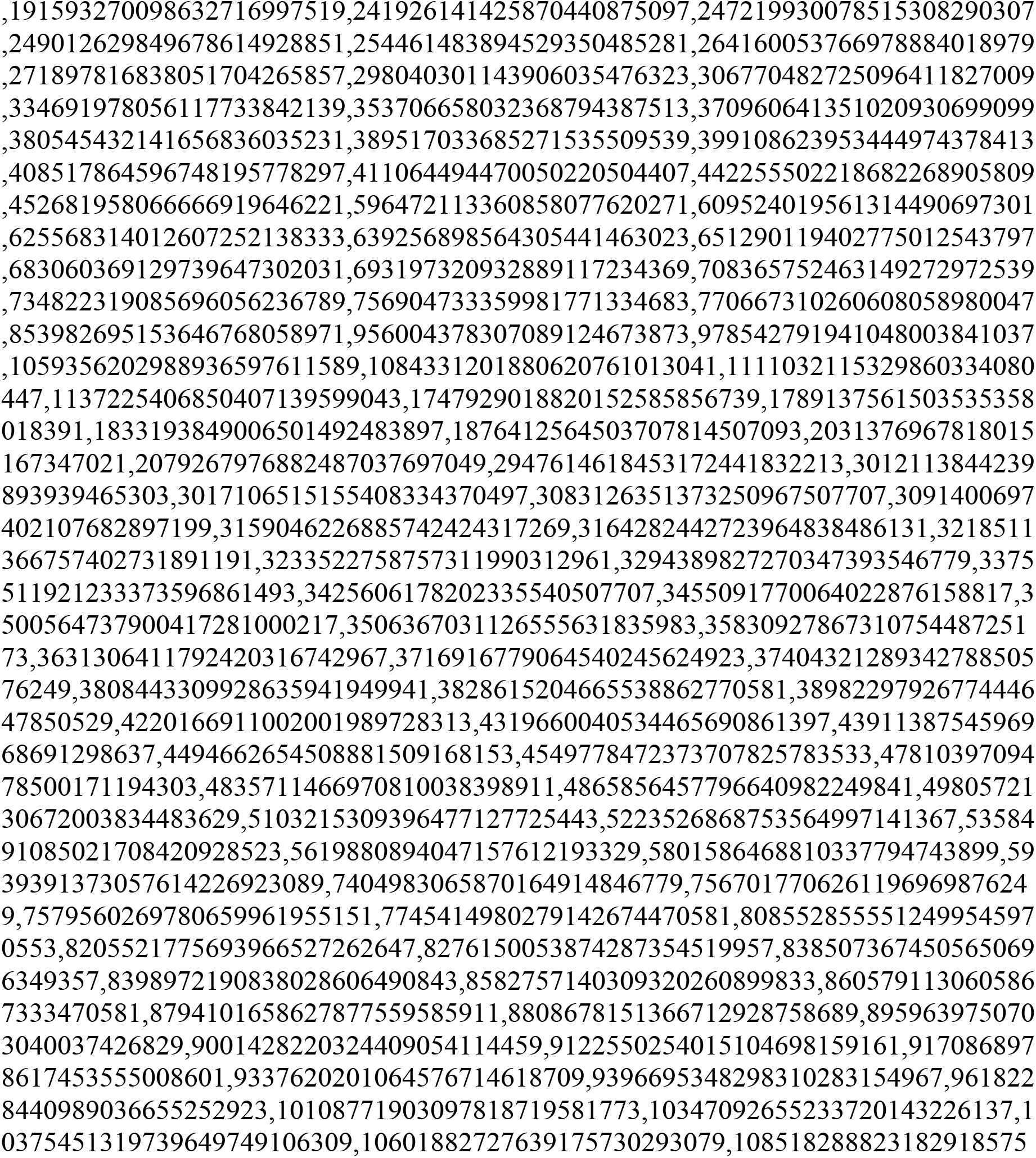
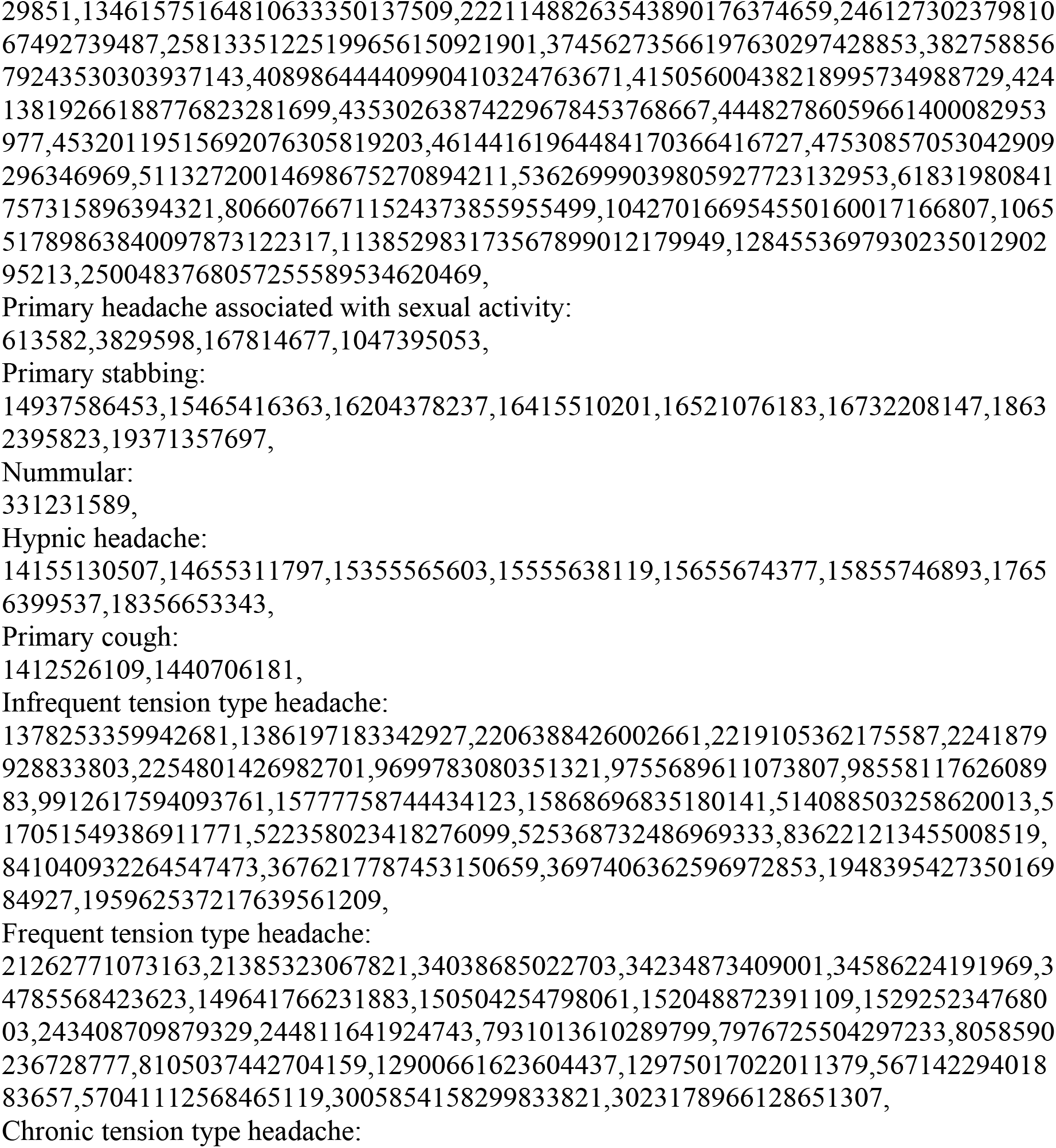
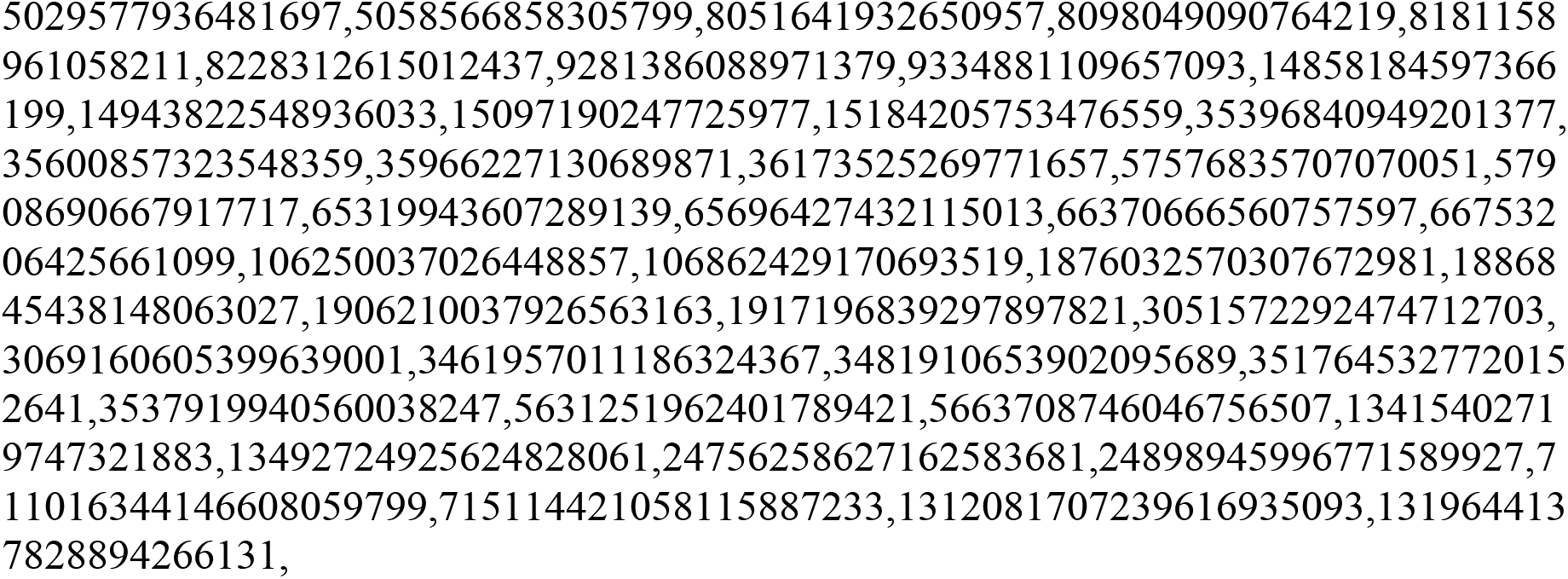
Prime number encodings for selected primary headache disorders:

We further generate, by hand, a list of mutually logically exclusive phenotypes (Table 3). This list in table 3 are then encoded as a list of composite numbers. We call this list L’.

**Table 3.**
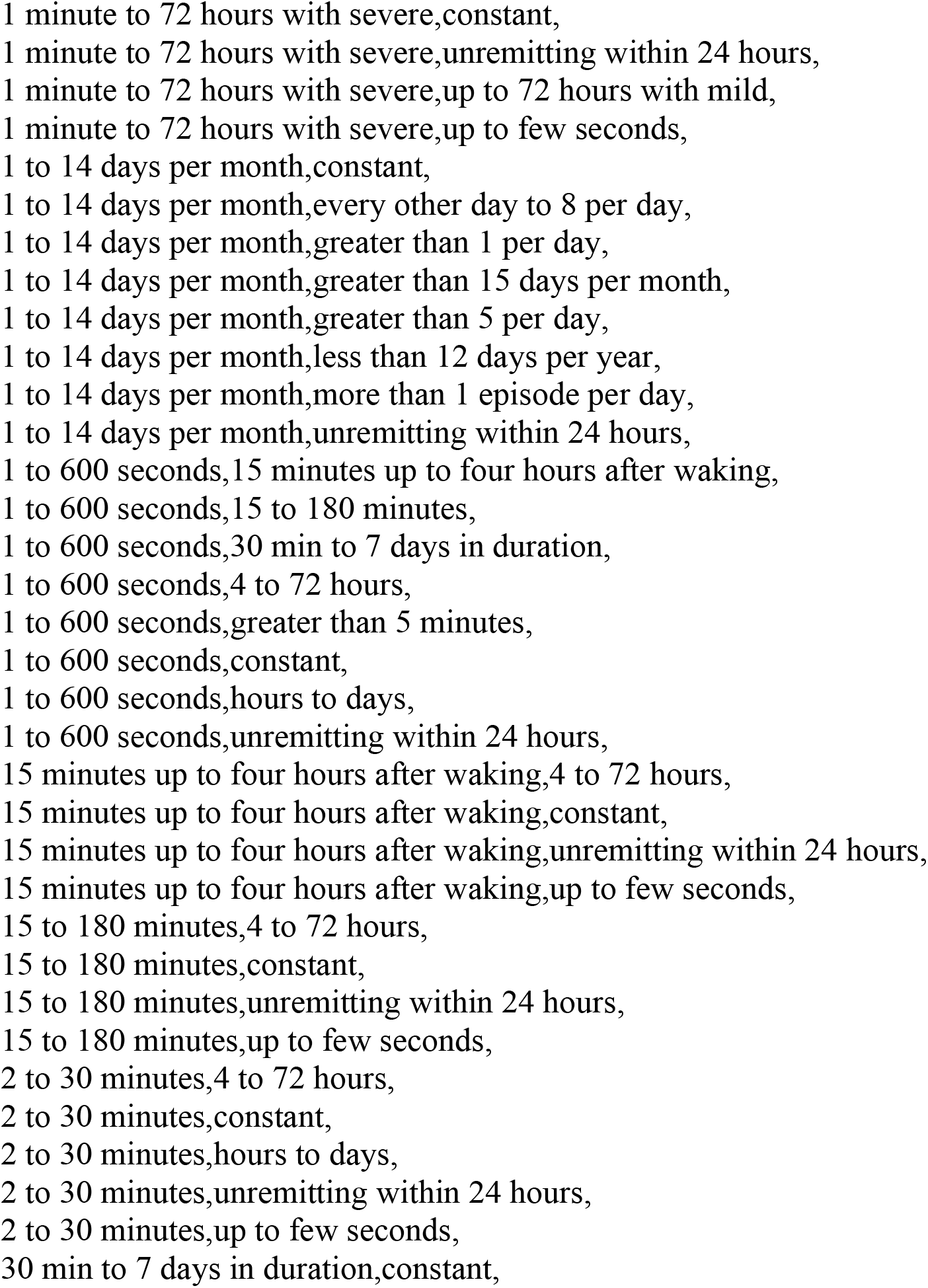
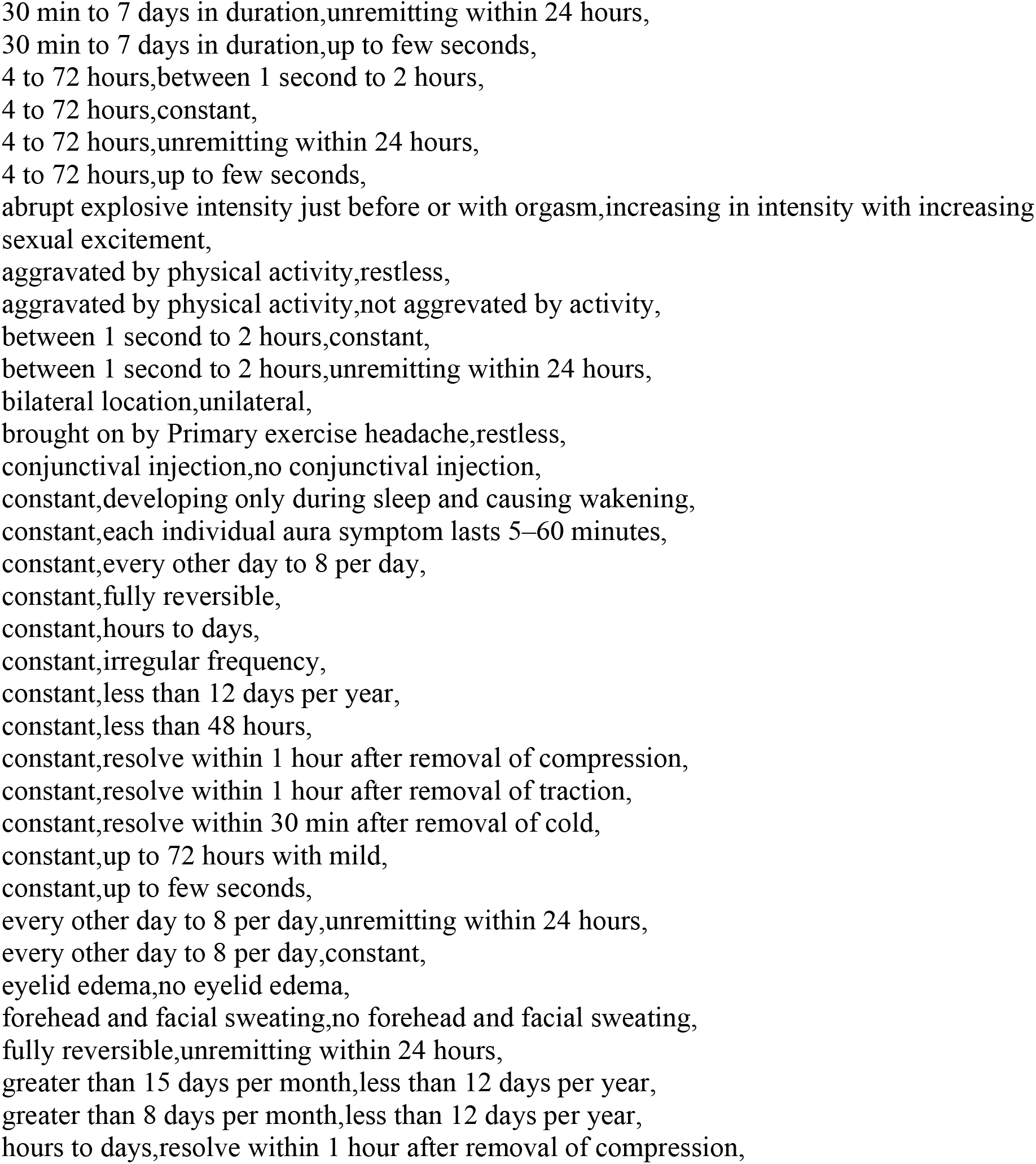
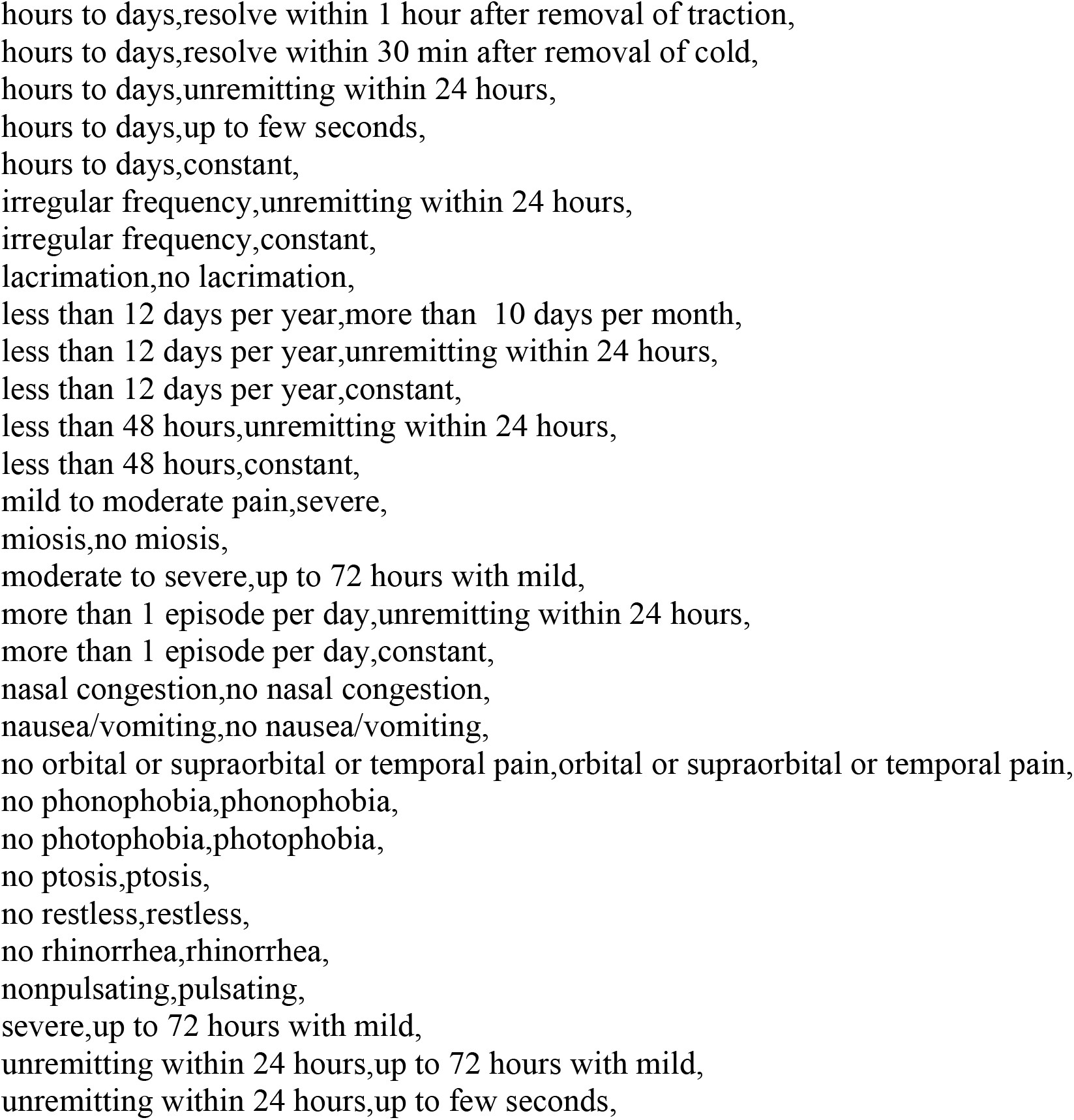
a list of logically contradicting phenotypes:

Any number in list M’ that is divisible by any number in list L’ are then excluded. The resulting list are therefore the totality of all possible patient clinical profiles with dual diagnosis that contains non-contradictory criteria. We then diagnosed these clinical profiles using the automated diagnosis technique discussed above. Since there are multiple duplicates when it comes to which dual diagnosis are possible; eliminating the duplicates generates the results presented in table 4.

**Table 4.**
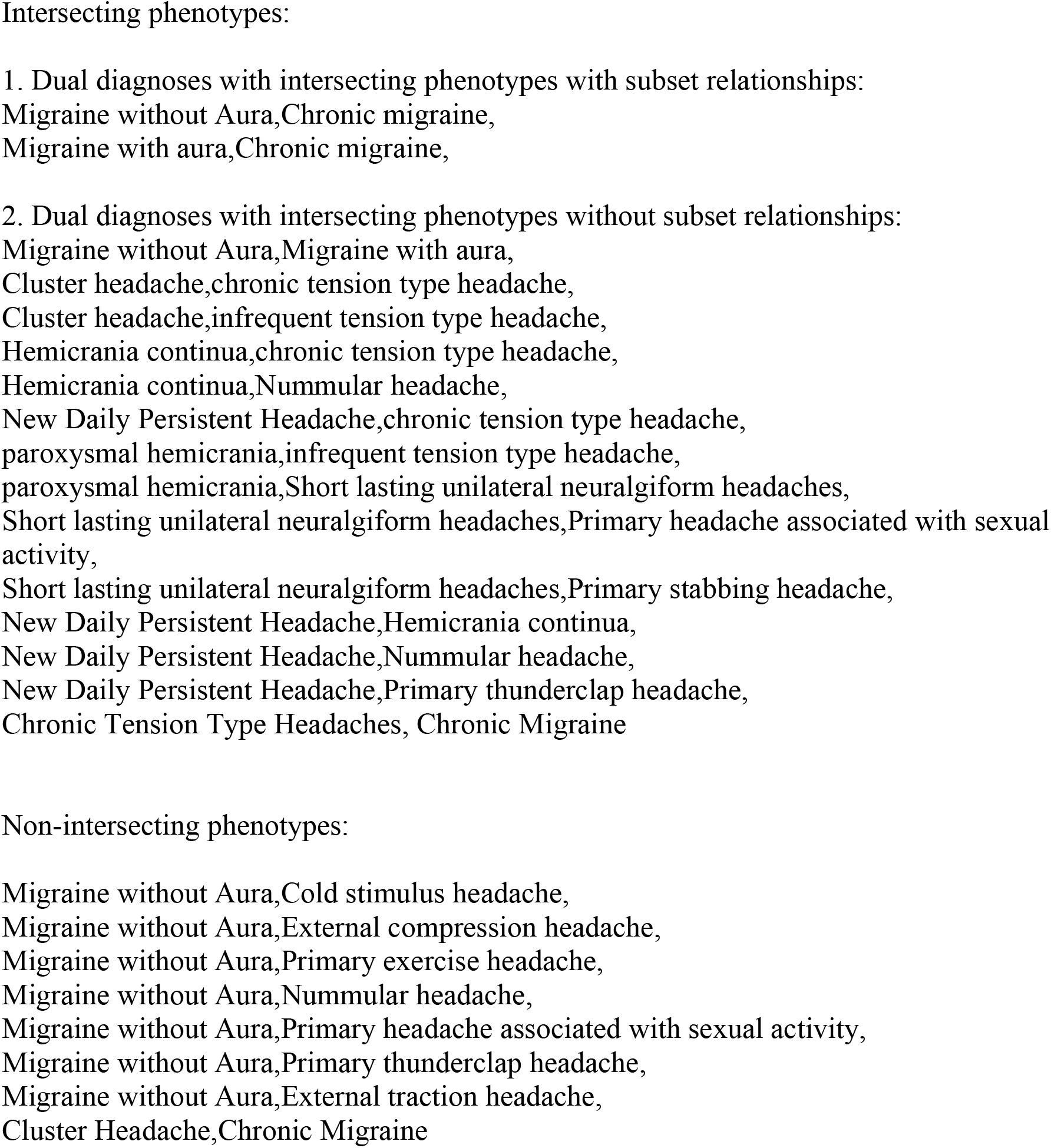
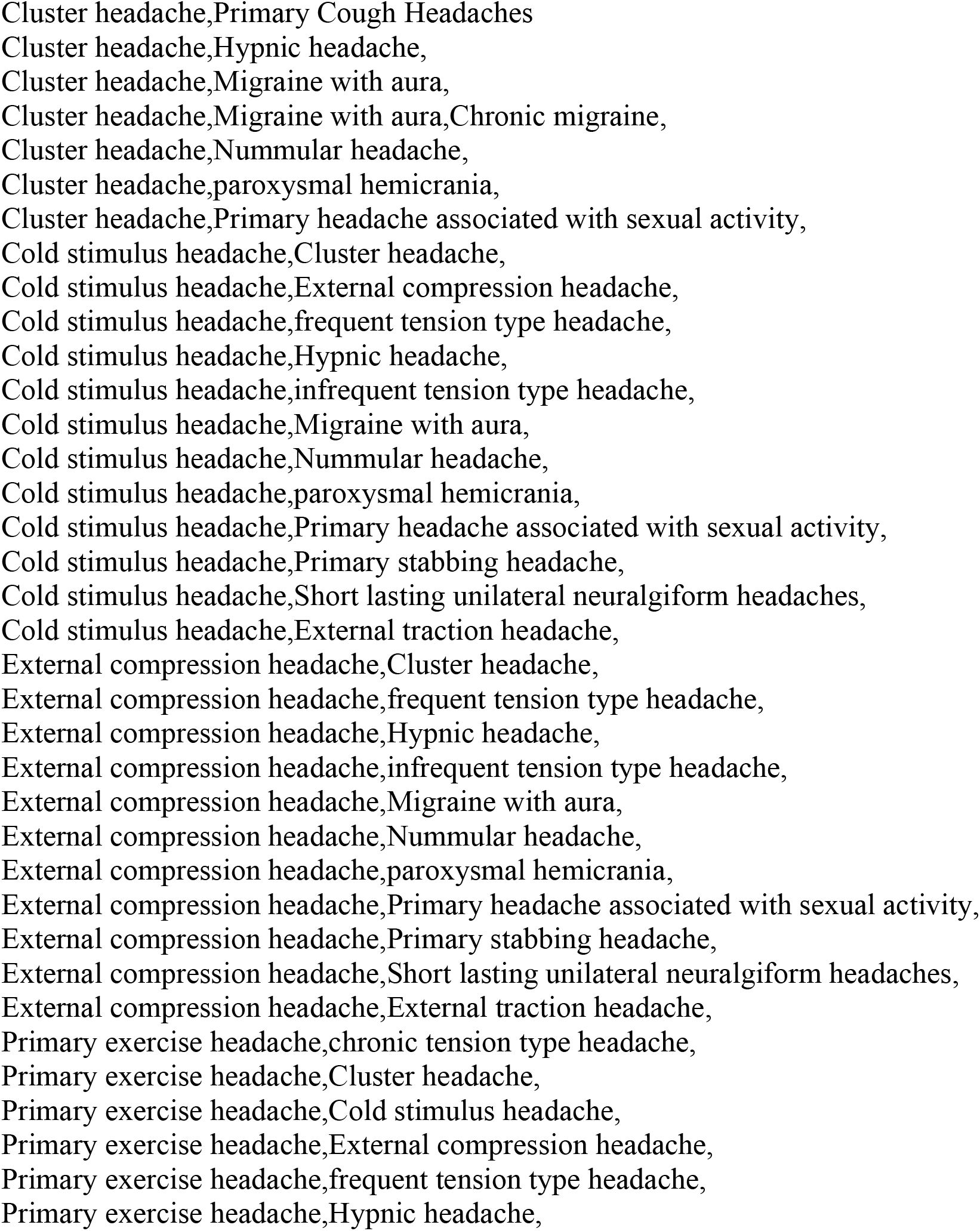
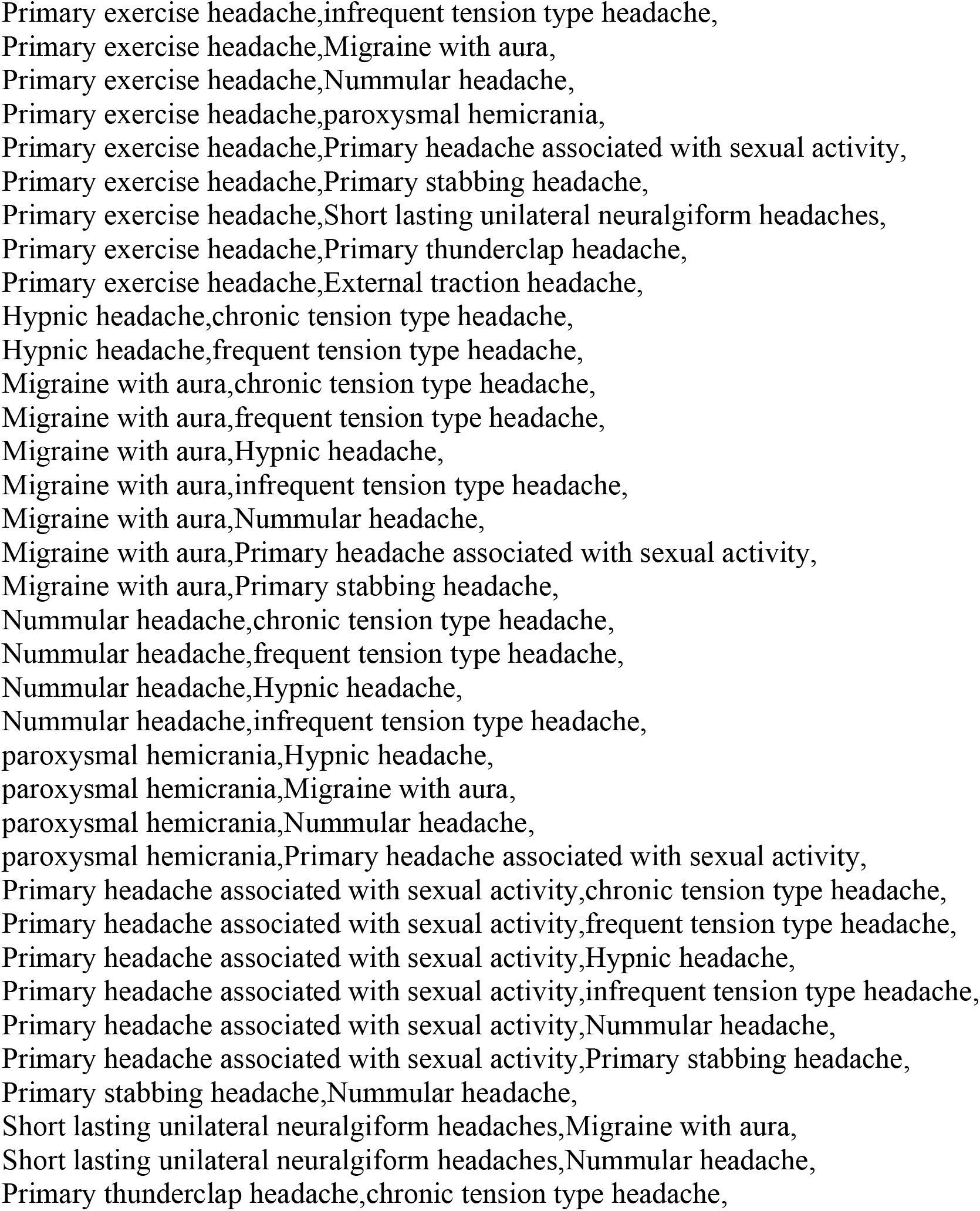
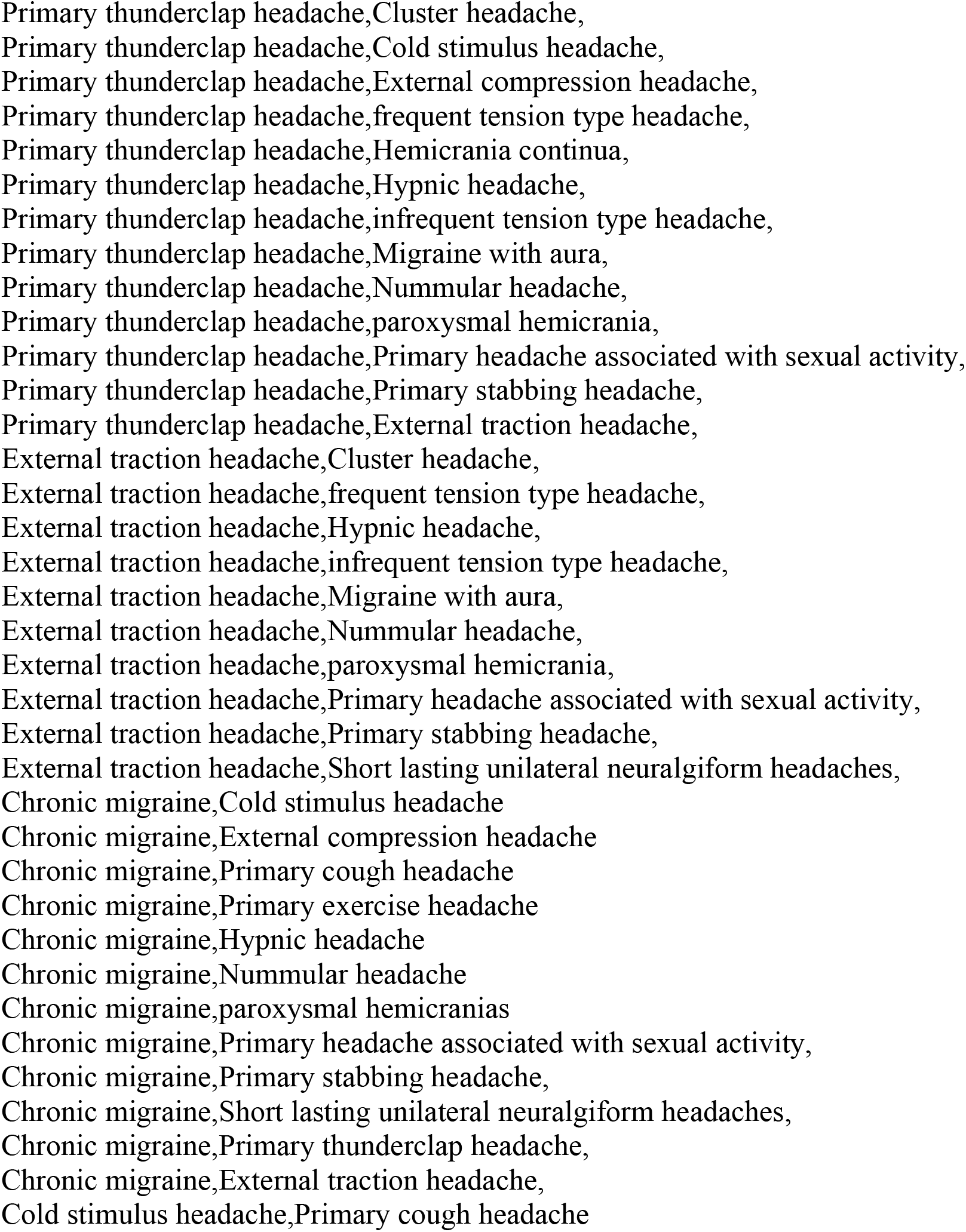
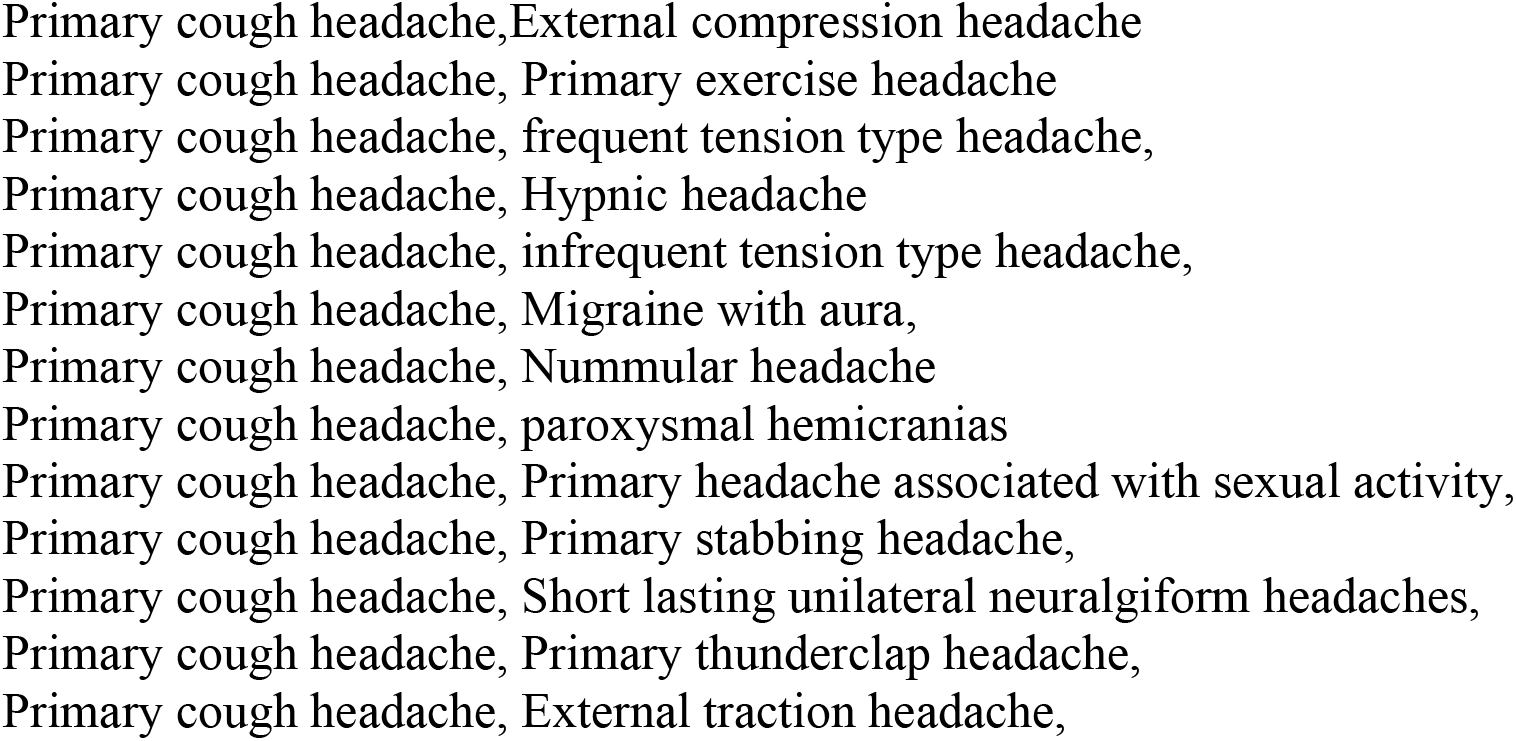
Logically possible dual-diagnoses, classified:

The above are implemented through custom codes written by the author in Haskell and are available for review upon inquiry.

### Analysis of results as “intersecting” vs “non-intersecting” phenotypes

We will now introduce a paradigm to interpret the results through the concept of “intersecting phenotypes”: Let us define a dual diagnosis pair as having “intersecting phenotype” if there exists at least one phenotype shared between the two diagnoses. If this does not exist then we describe the pair as having “non-intersecting phenotype”. (Non-intersecting phenotypes are “less interesting” in the sense that it is always possible to obtain dual diagnosis whose phenotypes are non-intersecting. We previously put forth this notion in a preprint.^12^) For example, primary stabbing headaches and SUNCT is a dual diagnoses pair with intersecting phenotype given that both contains duration measured in seconds. On the contrary, cold induce headache and nummular headaches is a dual diagnoses pair with non-intersecting phenotype as the phenotypes in their criteria do not overlap.

We can now classify our results as the following: first we separate those with intersecting phenotypes from those with non-intersecting phenotypes. We further differentiate those with intersecting phenotype due to a subset relationship versus those with intersecting phenotype without a subset relationship. For example, chronic migraine is a subset of migraine without aura since chronic migraine is defined as the existence of migraine without aura or migraine with aura greater than 15 days per month. This would be an example of an intersecting phenotype which contains a subset relationship.

## Results

A total of 103 prime numbers were used to encode phenotypes from the included ICHD3 criteria diagnosis. (Table 1) A total of 578 encodings were generated. (Table 2) A total of 99 pairs of illogical phenotypes were found. (Table 3) Once illogical phenotypes were excluded, a total of 253,842 composite numbers representing unique dual-diagnosis clinical profiles were obtained. A number of profiles, although unique, yield duplicate dual diagnoses; once these duplicates are removed, we obtained 145 possible logical dual diagnoses. (Table 4)

Using our classification schema above, we obtained the following: 2 dual diagnoses with intersecting phenotypes due to subset relationships, 14 dual diagnoses with intersecting phenotype without subset relationships. The remaining 129 dual diagnoses are results of non-intersecting phenotypes. This breakdown of results is incorporated in Table 4. The intersecting diagnosis that are subsets of each other contain only two pairs of dual diagnosis: migraine with or without aura and chronic migraine. This is to be expected given that we define chronic migraine through the existence of migraine with aura or migraine without aura.

## Discussion

In clinical practice, patients with two concurrent ICHD3 diagnoses are common: Indeed, when managing secondary headaches with etiology which cannot be treated, it is recommended that one treat the phenotype of the disease; some would even suggest that secondary headaches may be triggers of underlying primary headaches and therefore should be treated according to the primary headache it provokes.^13^ This is also the preferred method for treatment of post traumatic headaches.^14^ Therefore, secondary headaches with a primary headache phenotype are a rather non-controversial manifestation of the dual headache diagnosis problem.

Dual diagnosis in primary headaches, however, can be the source of controversies as discussed in our introduction. This stems from the fact the primary headache disorders are phenotypically defined; controversies arise not necessarily from a disagreement in regards to pathophysiology, but rather definition. In other words, the problem is a Kantian one: What are the conditions of possibilities of being diagnosed as a specific kind of primary headache disorders? What are the redundancies in the contemporary classifications of headache disorders? The results presented here may provide some answers. Our discussion will be organized around a few canonical diseases.

### The migraine/TTH Dilemma

While the split between migraine without aura and tension type headaches is an old one, that migraine with aura and tension type headache can be co-diagnosed is not surprising; migraine with aura patients can often have a headache that is not phenotypically like migraine without aura - the exemplary case being migraine with aura without headaches. More peculiar, however, is that it is actually possible to have migraine with aura diagnosed concurrently with chronic tension type headache and chronic migraine. This is due to the “loophole” that migraine with aura and chronic tension type headache can coexist. In this case, if aura occurs greater than 15 days per month, one can obtain these “monster” diagnoses that is the duo of chronic migraine, migraine with aura, chronic tension type headache.

### Cluster Headaches, TAC, and SUNCT/SUNA

Our results also highlight some of the peculiarities of our contemporary classification of TAC. Firstly, it is not surprising to notice that cluster headache can be diagnosed together with hypnic headache. It is also not surprising that cluster headache has a non intersecting phenotypical relationship with migraine with aura. This is, of course, a source of contemporary debate mentioned previously. (Indeed one can make the same argument for any TAC being co-diagnosed with migraine with aura following the same logic.) What is more surprising is that, following the loophole above, since cluster headache can be diagnosed together with migraine with aura, it is theoretically possible to have a chronic migraine form of cluster headache with migraine with aura: If a patient contains a co-diagnosis of migraine with aura with 15 days of aura and cluster headache, that person satisfies chronic migraine.

Our data also confirms the dilemma between paroxysmal hemicranias and cluster headaches in clinical diagnosis. Fortunately, an indomethacin trial can often be used to differentiate the two.^15^ Nevertheless, a co-diagnosis between cluster headache and paroxysmal hemicranias is possible as cluster headache is not defined as unresponsive to an indomethacin trial.

Short lasting unilateral neuralgia form headaches can be co-diagnosed with sex headache and primary stabbing headache. This is due to the fact that all of these headache types are short lasting in nature, and raises the theoretical possibility, of subtypes of short lasting unilateral neuralgia form headaches to be further classified by their provocating phenotypes.

### New Daily Persistent Headaches and Status Migrainosus

New daily persistent headache can be logically diagnosed with chronic tension type headache; empirically multiple researches in new daily persistent headache separate migraine versus tension type headaches phenotype in NDPH.^9^ However, according to the IC HD3 definition, it is actually impossible to diagnose new daily persistent headache concurrently with either migraine with or without aura. This is both due to 1) a limitation of the current study of not including status migrainosus and 2) from an artifact of the classification guideline which suggests that migraine, both migraine without aura as well as migraine with aura, must not be constant - migraine without aura is limited to 72 hour duration according to the guideline and migraine with aura also is limited by the fact that the migraine aura must be reversible. It is only the concept of status migrainosus which allows one to bridge this gap and therefore the possibility of migrainous NDPH.

The concurrent diagnosis of new daily persistent headache with hemicranias continua, nummular headache, or thunder clap headache essentially describes the subclassification of new daily persistent headache that was presented by Rosen in a prior review article.^16^ For instance, the lock sided headache nature of some new daily persistent headache may be attributed to a cervicogenic cause, of which nummular headache, being lock sided in nature, can be another manifestation. That new daily persistent headache can be concurrently diagnosed with thunderclap headache simply shows us that there is a subtype of new daily persistent headache which could be a persistent form of RCVS, as proposed by Rosen.^16,17^

### Non-intersecting phenotypes

Non-intersecting phenotypes represents the majority of dual diagnoses. This leads to the question of headache trigger in primary headache disorders. For example, it is possible to co-diagnose migraine without aura with cold induced headaches, compression headache, or exercise headache. It is also well documented that migraine can be triggered by cold (temperature), compression, or exercise.^18-21^ In the setting of these co-diagnosis then, is a dual diagnosis of migraine without aura with cold inducing headaches simply migraine without aura that’s being triggered by a cold weather? (Of note, it is actually hot rather than cold weather that triggers headaches more in Tanik’s study; this brings up the question of a theoretical possibility of “hot induced headaches”.) The same statement can be made regarding compression or exercise induced headaches.

While this may appear to be a trivial observation, its existence is an exploitation of a loophole in our current classification suggests to us that perhaps we ought to consider cold, exercise, compression as migraine subtypes when they co-exist with migraine or any other primary headache disorders. This notion is similar to Schankin’s suggestion that secondary headaches with primary headache diagnoses should be classified as such.

### Limitations

Despite our attempt to rule out obvious logical contradictions in the methods, this is not a fool-proof method. For example concurrent diagnosis of cold induced headache with hypnic headache is on paper logically consistent but in practice impossible - it would be unlikely for somebody to wake up at night from a hypnic headache while triggering that same headache through a cold stimuli. However, these sorts of occurrences in the data appears to be rare. The only other two instances following a similar argument for compression headache and hypnic headache pairing as well as the primary cough headache and hypnic headache pairing.

Finally, status migrainosus is considered a “complication of migraine” in the ICHD3 and therefore excluded based on our inclusion criteria. This proves to be an important omission as it limits the duration of migraine attacks. That status migrainosus’ complicated relationship to intractable headaches is not lost for those conducting research in status migrainosus.^22^ As such, although our omission causes the study to be incomplete, but shows us the importance of a rethinking of duration in migraine definitions.

## Conclusions

Prime number encoding of ICHD3 allows for an exhaustive study of the structure of contemporary headache diagnoses. In our pilot study of this approach, these important results are obtained: 1) Triggers may be used to delineate migraine subtypes. 2) Status migrainosus is required for modeling migraine intractability in our classification. 3) The possibility of dual diagnosis of chronic migraine and migraine with aura allows for loopholes in diagnoses which enable the construction of “monsters” in headache diagnosese such as co-diagnoses of chronic migraine, chronic tension type headaches, migraine with aura. This maybe undesirable in our classification.

## Data Availability

All data produced in the present work are contained in the manuscript

## Glossary

NDPH: New Daily Persistent Headache
ICHD3: International Classification of Headache Disorders, 3^rd^ Edition

## Article highlights

- Prime number representations of headache phenotypes allows for automated diagnosis based on ICHD3 criteria.
- Dual diagnoses can be classified and evaluated using the automated diagnosis methods above.

## Declaration of Interests

PZ has received honorarium from Lundbeck Biopharmaceuticals, Board Vitals, and Fieve Clinical Research. He collaborates with Headache Science Incorporated without receiving financial support. He has ownership interest in Cymbeline LLC.

## Study Funding

N/A.

## Acknowledgements

N/A.

## Notes

### Funding Statement

This study did not receive any funding

### Summary of Updates

A few data points are added and fixed; specifically in regards to co-diagnosis with chronic migraine and also primary cough headaches.

